# What do women want to see in a personalized breast cancer risk report? A qualitative study of Asian women of two countries

**DOI:** 10.1101/2023.08.29.23294527

**Authors:** Fatma Aldila, FJ Fiona Ng, Jessica Audrienna, Lim SJ Lynn, Shannon Tang, Sabrina Gabriel Tanu, Eric Aria Fernandez, Faustina Audrey Agatha, Marco Wijaya, Stevany Tiurma Br Sormin, Levana Sani, Astrid Irwanto, Samuel J. Haryono, Jingmei Li, Alexandre Chan, Mikael Hartman

**Author notes:** Corresponding author: (FA). These authors contributed equally to this work (Fatma Aldila, FJ Fiona Ng). These authors also contributed equally to this work (Faustina Audrey Agatha, Marco Wijaya, Stevany Tiurma Br Sormin).

## Abstract

**Introduction:** A breast cancer risk assessment tool using Polygenic Risk Score (PRS) of 313 single nucleotide polymorphisms and a risk prediction algorithm from the Gail Model had been established and validated for Asian populations. However, effective ways for delivering personalized risk information have not been fully explored yet. Through facilitated focus group discussions, we assessed the preferences of women from two Asian countries regarding the presentation of risk results, the level of detail provided for explanations, and recommendations for follow-up actions.

**Methods:** We conducted ten focus group discussions between July and October 2022 with thirty-two Asian women above the age of 25. The discussions aimed to evaluate the views and perceptions of women in Indonesia and Singapore in relation to personalized breast cancer risk assessment. All participants received either a detailed high-risk or low-risk mock report and were assigned to either high-risk result or low-risk result focus group based on the report they had randomly received. All focus group discussion content was then thematically analyzed.

**Results:** Participants indicated a preference for a comprehensive report that included follow-up steps and information for managing their breast cancer risk. Additionally, they highlighted the need for visuals without colors that project intimidation as well as a summary on the first page of the report to support interpretations. In the context of the report’s content, participants preferred personalized recommendations for risk reduction, and communication styles could be enhanced through the use of simplified language. Furthermore, we found out that Singaporeans receiving low-risk results were less likely to seek additional physician follow-ups compared to Indonesians, due to their greater breast cancer prevention awareness. Finally, participants found the report useful and would like to see similar reports on other diseases in the future.

**Conclusion:** Overall, most patients prefer a test report to strike a balance between content and complexity. The study also highlighted the importance of the psychological impact of patients receiving their test reports, which is greatly influenced by the patients’ degree of understanding and interpretation of the reports. Lastly, as most patients would likely increase their engagement with their physicians upon receiving their test results, future studies could be extended to physicians who are directly involved in the patient care delivery of breast cancer prevention.

## Introduction

Breast cancer (BC) is the most common cancer and the leading cause of cancer-related deaths among women. In 2020, there were 2.3 million new cases of women diagnosed with BC and 685 000 deaths worldwide [1]. In the past few years, there has been a rise in popularity of studies using polygenic risk scores (PRSs) to determine a person’s risk of BC [2–4] PRS, which sums the impact of multiple common genetic variants to estimate disease risk, has demonstrated the potential to improve the identification of individuals at high risk of developing BC [5].

Early diagnosis of BC through mammography screening has been shown to reduce deaths from the disease [6,7]. However, mammography does not come without risks. False positive tests and overdiagnosis of tumors that may never advance into a clinical stage that endangers life are commonly cited downsides of mammography screening [8,9]. In Asia, the benefits of mammography screening are further impeded by the lack of organized screening programs, and the fear of mammogram procedural pain [10–12].

Numerous risk predictors for BC, such as family history, classical BC risk factors, mammographic density, and PRS, thus play a pivotal role in determining which women are most likely to benefit from surveillance [13–16]. A BC risk assessment tool using PRS assessment of 313 single nucleotide polymorphisms and a risk prediction algorithm from the Gail Model had previously been established [17,18], and validated [19–21] for Asian populations. The generated risk report provides information on BC risk stratification and personalized risk-reducing recommendations, which will help doctors in delivering tailored care plans. To improve the communication and delivery of genetic based screening test reports, for example, studies were conducted in the United States [22] and the United Kingdom [23] to provide recommendations on the report format. However, the findings reported in the two studies may or may not be applicable to all clinical settings, especially in the Asian context and for BC in particular.

Beyond healthcare providers, the potential impact of disclosing the BC risk assessment report to its targeted customers, namely healthy women with no history of BC, is still not well understood. Almost all published studies are sourced from non-Asian countries where acceptance of BC risk assessment test could be different from the Indonesian and Singaporean populations. Hence, effective ways of returning personal risk information comprising predictions from various genetic and non-genetic risk factors remain yet to be explored. Through ten facilitated focus group discussions (FGDs), we assessed the preferences of thirty-two women from two Asian countries regarding the presentation of risk results, the level of detail provided for explanations, and recommendations for follow-up actions.

## Methods

This was a qualitative study involving focus groups. FGD was deemed most suitable as it allowed for a deeper discussion on the perception of healthy women in Indonesia and Singapore in relation to BC risk assessment testing. Indonesia and Singapore were chosen because the countries have different policies and practices of BC screening programs. The clinical BC screening program in Indonesia is opportunistically designed as a response to symptoms reported by women. Meanwhile, Singapore has adopted a nation-wide BC clinical screening program referred to as BreastScreen Singapore since 2002 [24,25].

The study was conducted in Indonesia from September to October 2022 and in Singapore from July to August 2022. The Standards for Reporting Qualitative Research (SRQR) was utilized as reporting guidelines. The checklist is shown in S1 File.

Ethical approval for the study was obtained from the Parkway Independent Ethics Committee of Singapore under approval number PIEC/2022/013 and Ethical Committee of Atma Jaya Catholic University of Indonesia under approval number 0007W/III/PPPE.PM.10.05/09/2022. Written informed consent was sought from all the participants before they were enrolled in the study.

### Eligibility criteria and recruitment

The inclusion criteria included women aged above 25 who had not been diagnosed with BC and were able to use the Zoom application.

Convenience sampling method was used both in Indonesia and Singapore. The study was promoted via social media platforms, such as Instagram and LinkedIn, online newsletters, and direct promotion to potential participants by the study team members. Online study promotion was selected because online marketing would be primarily used to advertise the product in the future. Interested individuals were asked to fill out the Microsoft Forms so that the study team members could contact them to check their eligibility and time availability. This form allowed the study team members to identify the individual participants’ information, including name, age, and an explanation of the study procedures was then provided to the eligible individuals, and informed consent was obtained via HelloSign. The participants were offered electronic money credits valued at Rp 50.000 in Indonesia or SGD 20 in Singapore as an incentive to participate. Participation and withdrawal were fully voluntary.

### Sample size calculation

Researchers aimed to arrange four different sessions of FGD in each country. Based on existing literature, data saturation of 90% or higher could be achieved by conducting at least four sessions of FGD [26,27].

### Development of focus groups and interview guide

Prior to the focus groups, all participants received either a detailed high-risk or low-risk mock report that is currently under development by NalaGenetics (S2 File). Apart from the detailed mock risk report, participants in Singapore also received a simplified version of the mock risk report (S4 File), which was adapted from an ongoing research program [28]. Both reports contained recommendations that help patients manage their risks and a personalized screening plan to identify the risk of BC early, improving the effectiveness of treatment. All participants were then assigned to either the low-risk result or high-risk result focus group based on the report they had randomly received. The decision to separate the participants into two cohorts was based on the assumption that people who receive high-risk results may have different opinions from those receiving low-risk results.

All FGDs were conducted via Zoom application. FGDs in Indonesia were conducted in Bahasa Indonesia by FA, while FGDs in Singapore were conducted in English by FN. Both facilitators were part of the investigation team, and they had both undergone training for FGD. FA also had previous experience moderating a few sessions of FGD.

Based on the FGD simulations that had been previously conducted, the maximum number of participants was capped at five per FGD session to give each participant enough time to answer a question before moving on to the next question. Participants were reminded that there were no right or wrong answers, allowing them to freely express their opinions. The questions asked during FGD covered aspects that were related to room for improvement in the report prototype. (S3 File). Confirmatory questions were asked following the closed-ended questions to avoid bias.

### Data collection

During the FGD, the facilitator began each session by introducing all members of the study team. Next, participants were reminded of the background and objectives of the study. An explanation and interpretation of the report were also provided. All discussions were digitally recorded using the recording feature via the Zoom application.

To ensure that the participants were actively engaged in the discussion, all participants were requested to turn on their web camera during their sessions. The facilitator aimed to finish each session within 90 minutes to prevent exhausting the participants, while ensuring that sufficient time was allocated to them before moving on to the next question.

### Data analysis

All audio recordings were manually transcribed verbatim and reviewed by two independent study team members for assurance. Participants’ identities were anonymized by assigning them a unique code. Language translation to English was performed manually for all FGD responses collected in Bahasa Indonesia.

Through an iterative comparison, inductive thematic analysis was performed. This analysis was performed by coding the FGD participants’ responses; similar codes were then subsequently grouped together to obtain sub-themes. Finally, relevant sub-themes were grouped together into broader themes. Any differences in opinions were resolved through a discussion among the study team members. No specific framework was used to assist with the analysis as the nature of the study was fully exploratory.

### Availability of Data and Materials

Permission to access patient data is restricted to the corresponding authors only. The shared information in this focus group comprises personal details that will not be made available for publicity due to ethical and legal reasons. Furthermore, anonymization of the complete transcript cannot be guaranteed due to a small group size.

## Results

Initially, a total of 64 individuals expressed their interest in participating in this study. Out of the 22 participants that were excluded from the recruitment, 2 were lost to follow up, 2 did not meet the inclusion criteria, 2 decided not to continue with the recruitment process, and 16 could not make it for the allocated time slot. This reduced the number of participants that were recruited to 42. Out of the 42 participants recruited, 10 participants did not manage to attend the FGD due to sudden work or other personal commitments. The final number of participants recruited from both countries was 32, with recruitment flow as shown in Figure 1.

**Fig 1.**
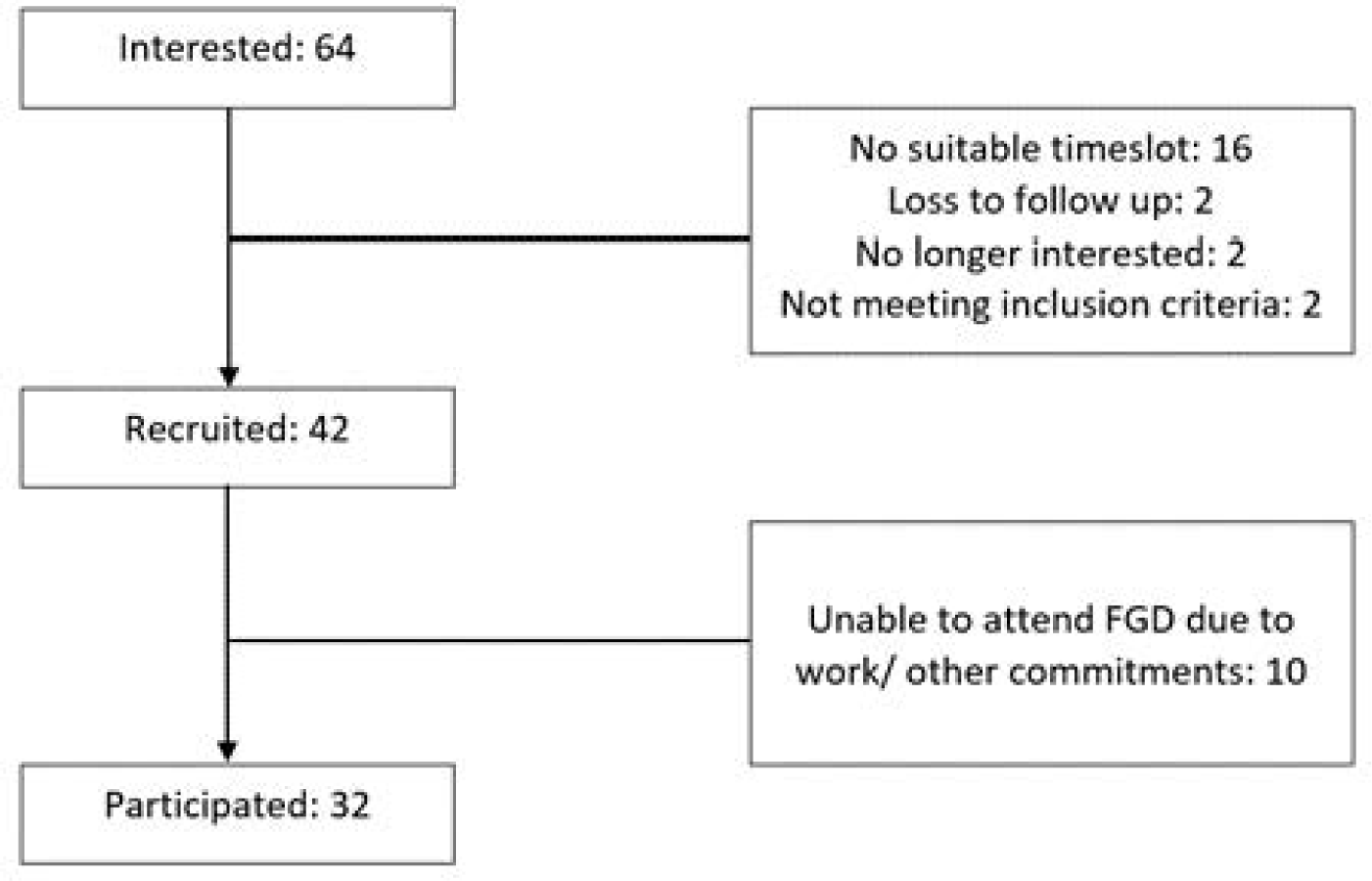
Flowchart of participant recruitment.

The demographics of the participants involved in the FGD in Indonesia and Singapore are summarized in Table 1. In Indonesia, participants were between 25 to 30 years old with a mean age of 27.9 years. Out of 15 total participants, 14 disclosed their ethnicity background. Eighty percent of the participants were Indonesian (n=12), and nine of them were from Jawa ethnic group. In Singapore, the age range of the participants involved was 25 to 49 years old with a mean age of 37.2 years. Eighty two percent of the participants were Chinese (n=14) while the rest were Indian (n=2) or Burmese (n=1).

**Table 1.** Participant demographics.

|  | Indonesia (n=15) | Singapore (n=17) |
| --- | --- | --- |
| Mean Age (years, range) | 27.9 (25-30) | 37.2 (25-49) |
| Ethnicity |  |  |
| Indonesia* | 12 (80.0%) | - |
| Chinese | 1 (6.7%) | 14 (82.3%) |
| Indian | - | 2 (11.8%) |
| Others/undisclosed | 2 (13.3%) | 1 (5.9%) |

\*Specific ethnic groups included: Jawa (n=9), Lampung (n=1), Minangkabau (n=1), Mixed (n=1)

A total of ten sessions of FGD were conducted for this study. Each session consisted of between 2 and 5 participants. The discussions with the low-risk result cohort were attended by a total of 18 participants divided into five different sessions. Similarly, the discussions with the high-risk result cohort were attended by a total of 14 participants split into five different sessions.

Four key themes were identified from the FGD sessions carried out. The first key theme gave insight on patients’ preference towards receiving a genetic risk report. The second key theme provided information around the areas of improvement for the report prototype (S2 File), and this included subthemes such as the content of the report, the design of the report, and the overall communication of the report. The third key theme provided insight into the potential impact of the BC risk assessment report on users. Meanwhile, the last key theme informed healthy women’s perceptions and expectations of BC risk assessment testing.

### Theme 1: More participants favor more information regarding their risk profiles

In Singapore, participants were asked to compare the detailed report against the simplified report (S4 File). 14 out of 17 participants (n=82%) would like to see reports that are more comprehensive in reporting their risk results and providing information on the follow-up steps to manage their BC risk.

> *“I prefer the one with more information … I feel that the other report is too short … [from the detailed report], you can find out which percentile you are [at] as compared to the general population … and I think there is more information with regards to the lifestyle changes [that] you can make.”* (Singapore FGD Participant 5, High-risk result report recipient)

Nonetheless, two of the Singapore FGD participants preferred the simplified report stated that a detailed report was not needed and would rather consult with their physicians if they needed additional information.

### Theme 2: Areas of improvement

#### Subtheme 2.1 The design of the report

In general, participants stated that the report exhibited an aesthetically pleasing and comprehensive visual presentation, characterized by an attractive design. Some participants also expressed a positive view of the report’s colorful and user-friendly format. However, a common observation among many participants was the difficulty in discerning important information, attributed to the use of light font color and a small font size. Additionally, a minority of participants raised concerns regarding the use of alarming color in a risk report, suggesting it might convey an unintended sense of alarm. Furthermore, there were suggestions to enhance the visibility of the test results in the report, with a preference for a concise summary of results on the first page of the report.

> *“In my opinion, the report [visual] is already good, [it is] also user-friendly in [providing] medical information … Yet, the colors [of the background and text] are not contrasting.”* (Indonesia FGD Participant 6, High-risk result report recipient)

> *“And also … like .. people normally focus on [text presented] in a larger font size, maybe that is from me … so that [the information] can be more visible.”* (Indonesia FGD Participant 12, Low-risk result report recipient)

> *“Report is comprehensive enough that it can give reassurance, but I am just wondering will there be another color that you all can choose other than pink? … where I am coming from is in terms of colors, it also brings across a message. If I give you a report that is red, of course you will be scared right?”* (Singapore FGD Participant 11, High-risk result report recipient)

#### Subtheme 2.2 The content of the report

##### A. Summary Risk Group

The majority of participants (n=24, 75%) found the summary of risk groups to be clear and were able to discern their combined risk group, as well as their categorization based on genetic and non-genetic risks. However, it is worth noting that a subset of participants (n=6, 19%) encountered challenges in comprehending their risk classification despite the presence of explanatory information.

> *“… [the information around risk group is] already clear, [the report shows] that the genetic risk [group] is low, the clinical risk [group] is also low … so there is overall elevated risk”* (Indonesia FGD Participant 7, High-risk result report recipient)

> *“I think the risk score is quite clear because they actually put a highlight for that.. I can tell whether I am in the high [risk] zone or the low [risk] zone …”* (Singapore FGD Participant 16, High-risk result report recipient)

##### B. Non-Genetic Risk

Twelve out of eighteen recipients of low-risk reports (67%) found the explanation of non-genetic risk calculation lacked clarity, especially concerning the use of medical terminology and the methodology employed for calculating the risk score. Furthermore, some participants sought clarification regarding the population data used in deriving the risk score. On the other hand, most participants (n=12, 86%) who received high-risk reports found the explanation to be sufficiently clear.

> *“So … there were only two [risk groups] for the non-genetic risk, right? Any score above 1.3 [%] would be classified into elevated [risk], right?”* (Indonesia FGD Participant 3, Low-risk result report recipient)

> *“The bottom part about this SEER Registry… I think as a lay person, I wouldn’t know what this is.”* (Singapore FGD Participant 7, Low-risk result report recipient)

##### C. Genetic Risk

Most participants (n=20, 63%) felt that the explanation of genetic risk calculation was clear, except for participants from the high-risk group in Indonesia. Overall, the provision of the graph was found to be useful as it aided participants in comprehending their respective risk groups.

> *“The genetic risk score is quite clear because it has indicated the percentage clearly as well as there is a graph … from the graph, I can differentiate what are the risk groups… and 5 years down the road, what will be the risk…”* (Singapore FGD Participant 4, High-risk result report recipient)

On the contrary, participants who felt that the genetic risk calculation was unclear found the supporting graph confusing, and they did not understand the technical language used.

> *“… since not all people understand statistics, the use of word ‘percentile’ would be confusing …next, for the graph [of genetic risk] … I understand [what the graph is about] but it took a while for me to discern …”* (Indonesia FGD Participant 10, Low-risk result report recipient)

> *“… next, for the graph [of genetic risk] … I do not really understand …”* (Indonesia FGD Participant 14, High-risk result report recipient)

##### D. Recommendations

Eighteen participants (56%) generally considered the recommendations provided in the report to be adequate and beneficial, while the remaining participants expressed concerns that these recommendations lacked specificity and were overly generic. Many participants (n=16, 50%) expressed a preference for more personalized recommendations. Additionally, there were suggestions to include easily understandable and reputable websites as supporting evidence for these recommendations.

> *“For myself, the lifestyle [recommendations] section is mostly clear, but it is too general so patients may be questioning what foods should be eaten less or more …”* (Indonesia FGD Participant 15, High-risk result report recipient)

> *“I feel that this is too generic, it’s not personalized for me … I feel that this is something I can just get from the internet.”* (Singapore FGD Participant 3, Low-risk result report recipient)

> *“Instead of having link to evidence, you could direct it to where I can find more information and where I can read more about it … because if I have a risk [of a certain condition] or I have this condition, I will immediately go and google … and you will read all the scary things [on the internet] which might not even be credible or applicable to you.”* (Singapore FGD Participant 9, Low-risk result report recipient)

#### Subtheme 2.3 Overall communication of the report

Study participants provided feedback aimed at enhancing the overall comprehensibility of the report. Suggestions included the incorporation of a glossary of medical terminology, which could prove beneficial for individuals without a background in medicine or science. Furthermore, participants recommended thorough proofreading to ensure the clarity and accuracy of the information presented in the report.

> *“… [the report] may need a glossary [summarizing medical terminologies], I think, as mentioned by Indonesia FGD Participant 1.”* (Indonesia FGD Participant 2, Low-risk result report recipient)

### Theme 3: Potential impacts of the BC risk assessment report

There were polarized reactions on how the participants would feel if the report prototype shown to them was really theirs. Negative emotions were shown by those receiving high-risk reports. Almost all participants (n=13, 92%) receiving high-risk reports said they would feel afraid, shocked, surprised, sad, worried, and anxious. However, there was one participant who would not feel shocked because she was aware that she had a family history of BC. On the contrary, most of the participants (n=13, 72%) who received low-risk reports mentioned that they were relieved. Some of them also reported that they would not be negligent in maintaining a healthy lifestyle because the risk score may change over time.

> *“[If this report was really mine], I would feel afraid, for sure. Because I have past experience of losing my grandmother (to breast cancer). But, regardless, I will focus on the treatment [to reduce the risk] and the steps I could take to address this.”* (Indonesia FGD Participant 7, High-risk result report recipient)

> *“If I were the one who receives the report, I would automatically feel relieved. Even though the risk in the future will still exist and may change [to become higher] …”* (Indonesia FGD Participant 4, Low-risk result report recipient)

> *“I think I will be very worried if I receive a report that says my risk is high … because one of my aunties has breast cancer …”* (Singapore FGD Participant 5, High-risk result report recipient)

Overall, most participants (n=12, 86%) receiving high-risk results reported that there would be a positive change in the relationship with their physicians as they would consult and follow-up with their physicians more often. There were also participants (n=4, 28.6%) who highlighted that the change in relationship with their physicians would depend on the understanding of the physician towards the report and the suggested interventions.

Meanwhile, among all study participants in Indonesia who received low-risk results, there was a consensus to engage in more frequent consultations with qualified physicians to ensure their risk remained low. Conversely, in Singapore, all study participants receiving low-risk results reported that there would be no changes in the relationship with their physicians.

> *“ … [I will be] consulting [with physician] more often; consulting [with physician] what kind of lifestyle is recommended, what would happen if I skipped [healthy eating and exercise] and cheated [from the recommendations] … whether those [nonadherent] things could increase the risk or not. I will ask more questions [to the physician].”* (Indonesia FGD Participant 15, High-risk result report recipient)

> *“Since we are in the low-risk group, there is no need to go and see my doctor asking about the report …”* (Singapore FGD Participant 9, Low-risk result report recipient)

Participants from Singapore and Indonesia had diverse questions for their physicians. When it came to monitoring and screening for BC, participants from Indonesia were curious about the advantages and potential drawbacks of mammography, whereas individuals from Singapore were inclined to compare it to the guidelines established by the Ministry of Health (MOH) in Singapore. In terms of lifestyle changes, participants were keen to inquire about dietary adjustments, including which foods to increase or decrease, as well as recommended physical activities. Questions also arose about prescribed medications and the significance of surgical procedures. Additionally, a small number of participants expressed interest in learning the correct method for conducting breast self-examinations.

> *“… what I would ask to my physician is on the type of mammography [examination], whether it has [negative] effect to me [or not]?”* (Indonesia FGD Participant 4, Low-risk result report recipient)

> *“I don’t know if your report is tailored to 40 - 49 years old … Compared to MOH recommendations, people over 50 [years old] don’t have to go [for mammography] every year … but this is not on your report …”* (Singapore FGD Participant 7, Low-risk result report recipient)

> *“Next, for the surgical [recommendation], [I may discuss] this later with a genetic counselor, whether we who are at the [high] risk should undergo surgery or not, maybe something like that, [so the question would be more on taking] a surgery [as an] option.”* (Indonesia FGD Participant 7, High-risk result report recipient)

> *“For therapy, [I want to know] more details about the risks… What should be done to reduce the risk? … and more details about the therapy…”* (Singapore FGD Participant 4, High-risk result report recipient)”

> *“Next, second, maybe about the self-breast exam, I may ask more or confirm [with my physician] about the exam, and how frequent I should do that.”* (Indonesia FGD Participant 12, Low-risk result report recipient)

Besides their physician, the participants would want to share and discuss the result of the genetic test with other parties. Most participants preferred sharing and discussing the report with family members or friends, especially those with medical backgrounds or who have been previously diagnosed with breast cancer.

> *“With whom I would talk to other than my physician if get this report? Perhaps, to my close friends or whoever [I know] who has ever been diagnosed with the same disease, if there are any. If not, I would only discuss this with my physician or the medical professionals [who take care of me].”* (Indonesia FGD Participant 4, Low-risk result report recipient)

> *“I will probably want to look at this report myself [and] do some research on my own … then probably talk to a family member then a doctor …”* (Singapore FGD Participant 6, High-risk result report recipient)”

The report also had the potential to change the participants’ outlook on their personal health. All the participants receiving high-risk results felt that the report would help increase their awareness of BC. They were also likely to change their lifestyle by following the recommendations given. Similarly, most of the participants receiving low-risk results stated that the report would change the way they view their health, their awareness of BC would also increase, and they would be more mindful of taking preventive actions.

> *“Yes, perhaps, [I would] be more aware, for example, that I would need to do self-breast exam more regularly. Generally, I would be more aware [of the breast cancer risk] due to the breast cancer risk prediction report I receive.”* (Indonesia FGD Participant 12, Low-risk result report recipient)

> *“Since I know I am in the high-risk group, I have to [know] what are the preventive methods I can take and moving forward … how do I minimize the risk to myself as much as I can.”* (Singapore FGD Participant 6, High-risk result report recipient)

### Theme 4: Future Disease Risk Assessment

The majority of the study participants found the report useful and expressed enthusiasm about receiving risk assessment reports for other diseases in the future. Suggestions from participants included exploring diseases such as cervical cancer, diabetes, and cardiovascular conditions.

> *“[I want to see a similar report for] types of diseases that are related to cancer. For example, cervical cancer, and maybe [risk assessment testing] related to fertility.”* (Indonesia FGD Participant 14, High-risk result report recipient)

> *“I am just thinking about the most common fields … people usually get diabetes, cancers…”* (Singapore FGD Participant 11, High-risk result report recipient)

## Discussion

To our knowledge, this FGD is one of the first studies conducted in two countries, Indonesia and Singapore, to evaluate the perception of healthy female participants towards testing results from personalized risk assessments for BC. Through the FGD, three areas of improvement for the detailed mock risk report were identified, which included the design, content, and communication styles of the report. These three areas coincide with recommendations by Farmer et al. [23] to improve understanding of patients and physicians to genetic test reports.

First, regarding the design of the report, there was feedback from FGD participants to avoid alarming font color to make the result less intimidating. Some sections of the report were also reported to be overly crowded with text. Similar findings were reported by Farmer et al. [23], where the patients recommended avoiding dense blocks of text and using different colors to differentiate ‘positive’ or ‘negative’ results.

Next, on the report content, this FGD found out that participants also demanded more information on recommended actions they could take to reduce their risk of getting BC other than improving the clarity of genetic and non-genetic risk information in the report. These correspond with a study by Huang et al. [29] that found out that one of the three things that affects people’s decision to undergo predictive genetic testing is the availability of treatment and prevention for the disease. The presence of recommended actions patients should take and resources that enable them to access further information and support for the disease are what patients are looking forward to in genetic testing reports [23].

Lastly, the report should be communicated in a style that is understandable to patients from non-healthcare professions. Not all patients understand medical or statistical jargon. Therefore, the addition of a glossary could be very helpful for patients. Even though the report is not meant to be self-interpreted by patients themselves without consulting with clinicians, making sure that patients understand the results of their risk assessment test is still imperative. Furthermore, having a user-friendly report could help to improve communications between the patients and their clinicians, thus allowing the counselling session to be more effective [22]. Patients who do not clearly understand their test results tend to show lower engagement in their personalized care [30] and are unable to adhere to recommended actions [31]. As a result, they are more likely to fail in reducing or maintaining their risk level.

A limitation in the study pertained to the utilization of mock reports, potentially leading to participants’ responses not accurately reflecting their genuine emotions. Participants may have reacted differently if real reports had been used, especially for those assigned to high-risk groups. However, the use of mock reports is a common practice in the field, employed to effectively survey a large number of participants.

The insights obtained from our study are useful to optimize the delivery of personalized risk assessment for BC. Most participants prefer a report that gives comprehensive information, rather than only informing them of which risk group they belong to. This is evident by the fact that the majority of FGD participants in Singapore preferred the detailed report. Patients’ preference over comprehensive genetic testing reports is supported by a study by Brewer et al. [32]. In addition to risk group, participants of the study could better understand their risk of getting BC recurrence when they were given a report that clearly showed the risk score, brief information to interpret the report as well as the graph depicting their position in the risk continuum.

The FGD identified the potential impacts of disclosing BC risk reports to patients. Patients would feel anxious upon receiving high-risk results and relieved upon receiving low-risk results from the testing. These findings correspond to the Singapore study conducted by Liow et al. [33], where the participants reported to be living in fear if they learned that their risk of acquiring BC was high. Given the nature of the testing as a diagnostic prediction, a negative psychological impact could occur following the risk assessment testing regardless of the test result. However, the anxiety level could be modest if patients understood what their report said [34] and the testing was performed by expert teams [35]. This emphasizes the importance of providing proactive personalized, pre- and post-test counselling conducted by counsellor who has sufficient understanding in personalized risk assessment for BC. Although many testing providers are now offering the option of delivering predictive genetic testing results via telephone or letter for convenience, participants may value the opportunity of having an individualized counselling session with the counsellor [36].

Our study found a key difference between Indonesian and Singaporean participants in our FGD. With greater knowledge and awareness towards BC prevention among the Singaporeans, it was observed that Singaporean participants receiving low-risk results would not want to have additional follow-ups with their physicians, compared to the Indonesian participants. Singaporean women may have a higher awareness level of BC screening due to the presence of clinical BC screening subsidies that allow women over 50 years to undergo mammography every two years at a more affordable cost [37]. However, the same initiative is absent in Indonesia, and high cost has been reported to be one of the main barriers to access BC screening by Indonesians [38]. This was also demonstrated by the disparity in understanding expressed towards mammography, with participants in Indonesia being unfamiliar with the procedure. With the inclusion of perspectives from two different countries of diverse ethnic groups that allowed for a more comprehensive study, we are confident that the results of this study could be broadly applicable to other Asian populations that share sociocultural similarities with Indonesia and Singapore.

There is no universally accepted guideline for test providers to ensure that their risk assessment report is understandable by patients. Results from the Gail Model may not be easily understood by patients [39,40]. Moreover, the format of reporting may impact the translation of genetic test results into clinical recommendations, the interpretation of test results, and ultimately, patients’ adherence to the recommended actions [41,42]. Accordingly, it is recommended that test providers that are planning to market their risk assessment services conduct studies to gain feedback from potential customers and identify potential impacts resulting from service that is still under development. Patients or healthcare service customers could be involved in the planning, development, and improvement of healthcare services [43,44]. The World Health Organization highly recommends patient engagement [45] to ensure that the delivered healthcare services result in maximum benefits and minimum risks for targeted populations.

Studies aiming to seek feedback on the report from physicians are also warranted. Participants, particularly those receiving high-risk results, would also consult more frequently with their physician in the hope of reducing their risk level. They might consider seeing another physician if the physician they are seeing is unable to give appropriate recommendations to act on their risk assessment report. This is imperative to make sure that the physicians, be it specialists or non-specialists, can communicate the report clearly and discuss a personalized care plan with their patients more comfortably. In addition, the FGD informed that the majority of the study participants found the risk assessment report to be useful and were interested in seeing similar reports for other diseases, including cancer, diabetes, and cardiovascular conditions. These findings shed light on the other diseases patients demanded which test providers could focus on in the future.

## Conclusion

This study summarized how a BC risk assessment report can be personalized for each individual to facilitate effective delivery of care plans for patients with different genetic and non-genetic risk profiles. With the increase in accessibility and availability of BC risk assessment service, many patients are interested in finding out more about their test results. Overall, most patients would prefer a test report to be well-balanced between content and complexity. The study also highlighted the importance of the psychological impact of patients receiving their test report, which is greatly influenced by the patients’ degree of understanding and interpretation of the reports. In our study, we discovered that Singaporean participants, who had higher awareness of breast cancer prevention, were less inclined to seek additional physician follow-ups after receiving low-risk results compared to Indonesian participants. Finally, as most patients would likely increase their engagement with their physicians upon receiving their test results, future studies could be extended to physicians who are directly involved in the patient care delivery of BC prevention.

## Data Availability

All data produced in the present study are available upon reasonable request to the authors.

## Acknowledgements

We would like to thank Professor Irwanto of Universitas Katolik Indonesia Atma Jaya for his guidance and training in the conduct and data analysis of focus group discussions.

## Supporting information

**S1 File. SRQR Checklist**

**S2 File. Detailed Report Prototype**

**S3 File. FGD Question List**

**S4 File. Simplified Report Prototype**

