## Supplementary material for "What do women want to see in a personalized breast cancer risk report? A qualitative study of Asian women of two countries": S2 File

**S2 File. Detailed Report Prototype**

**English Version**

Low Risk Report ([Link](https://nalagenetics.sharepoint.com/:b:/r/sites/clinical-studies/Shared%20Documents/General/%5BID%5D%20Clinical%20Studies%20Documents/PERCEPTION%20-%20Nalagenetics%20x%20MRCCC%20x%20SJH%20Initiatives%20-%20Breast%20Cancer%20RPS%20FGD/Dummy%20Reports/SG%20Dummy%20Report%20Low%20Edited%202.pdf?csf=1&web=1&e=NJruy9))
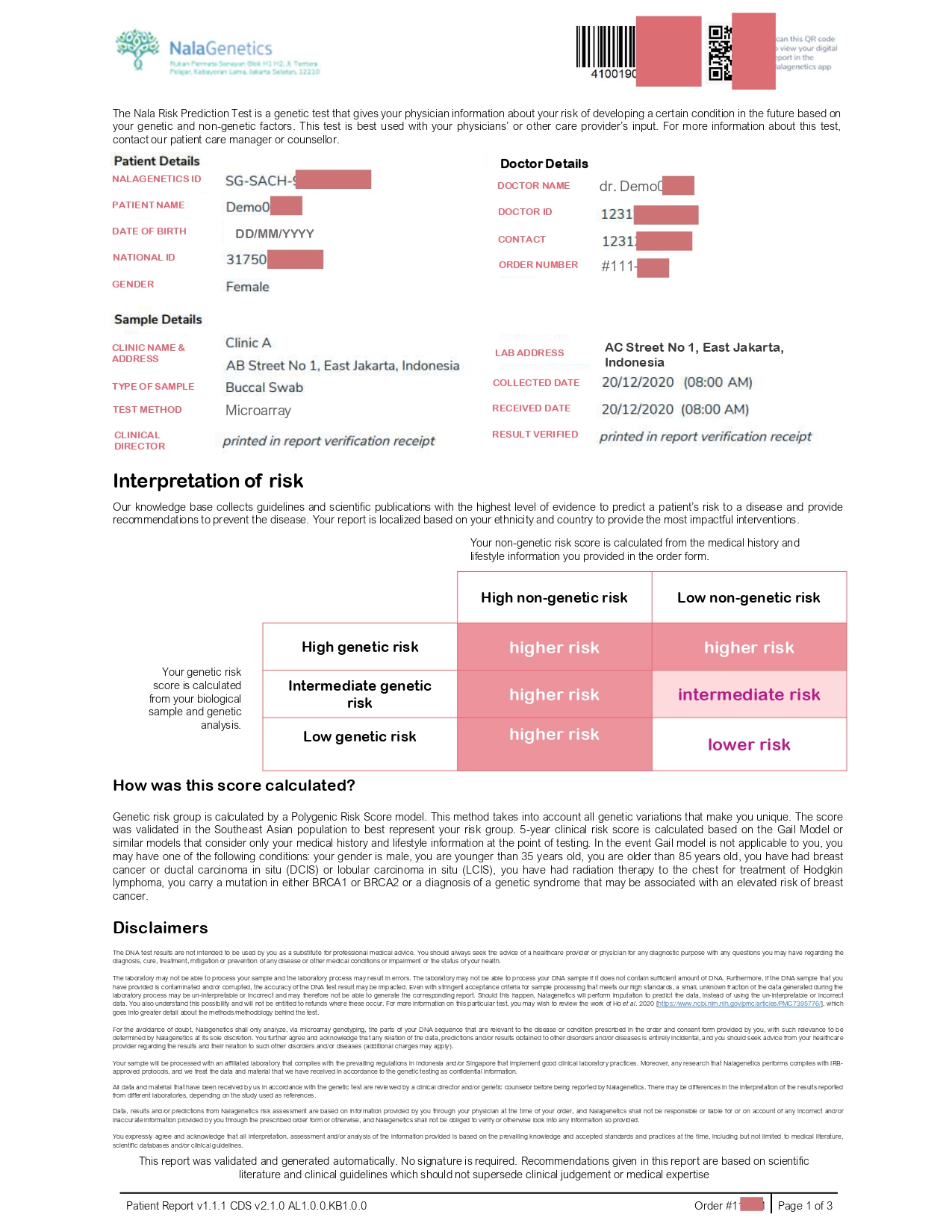

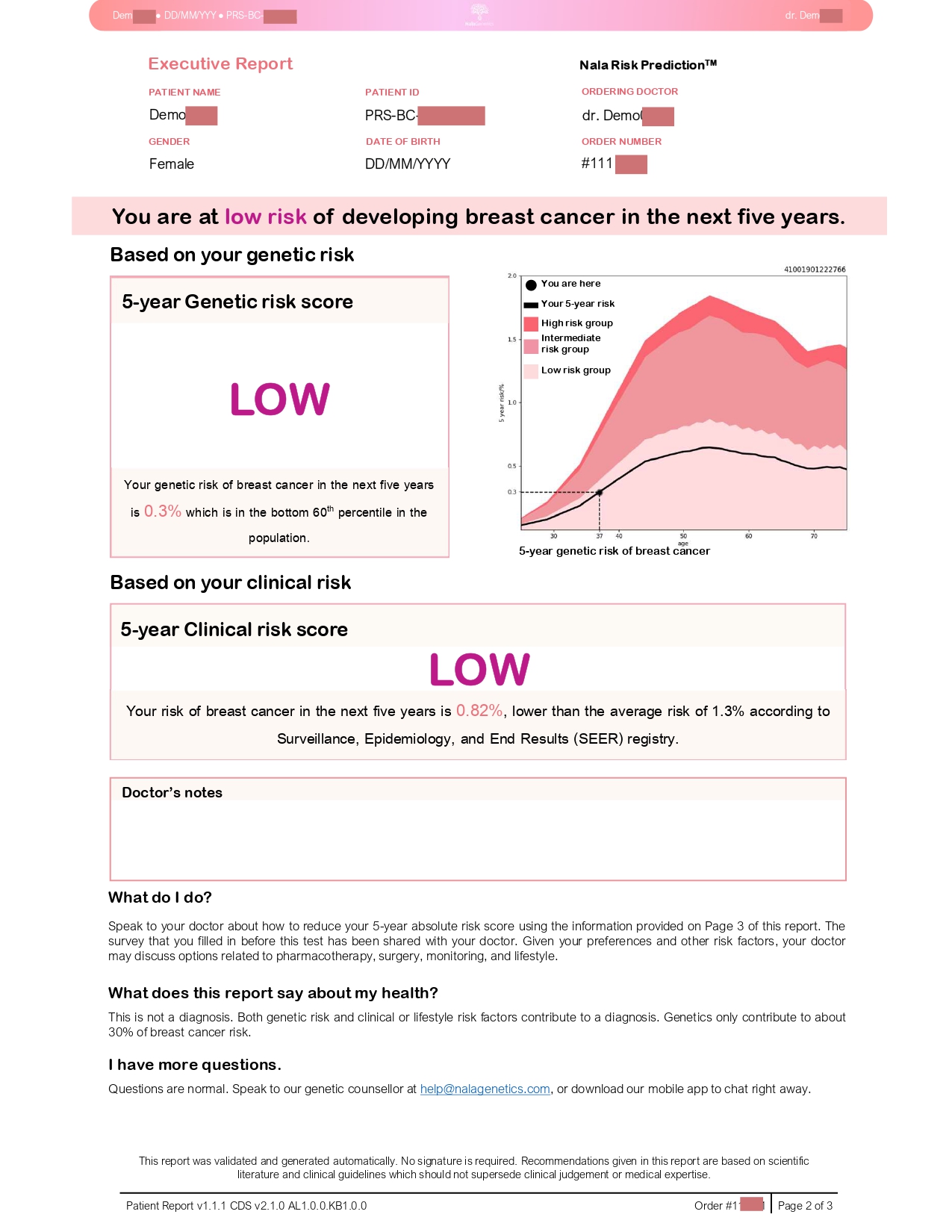

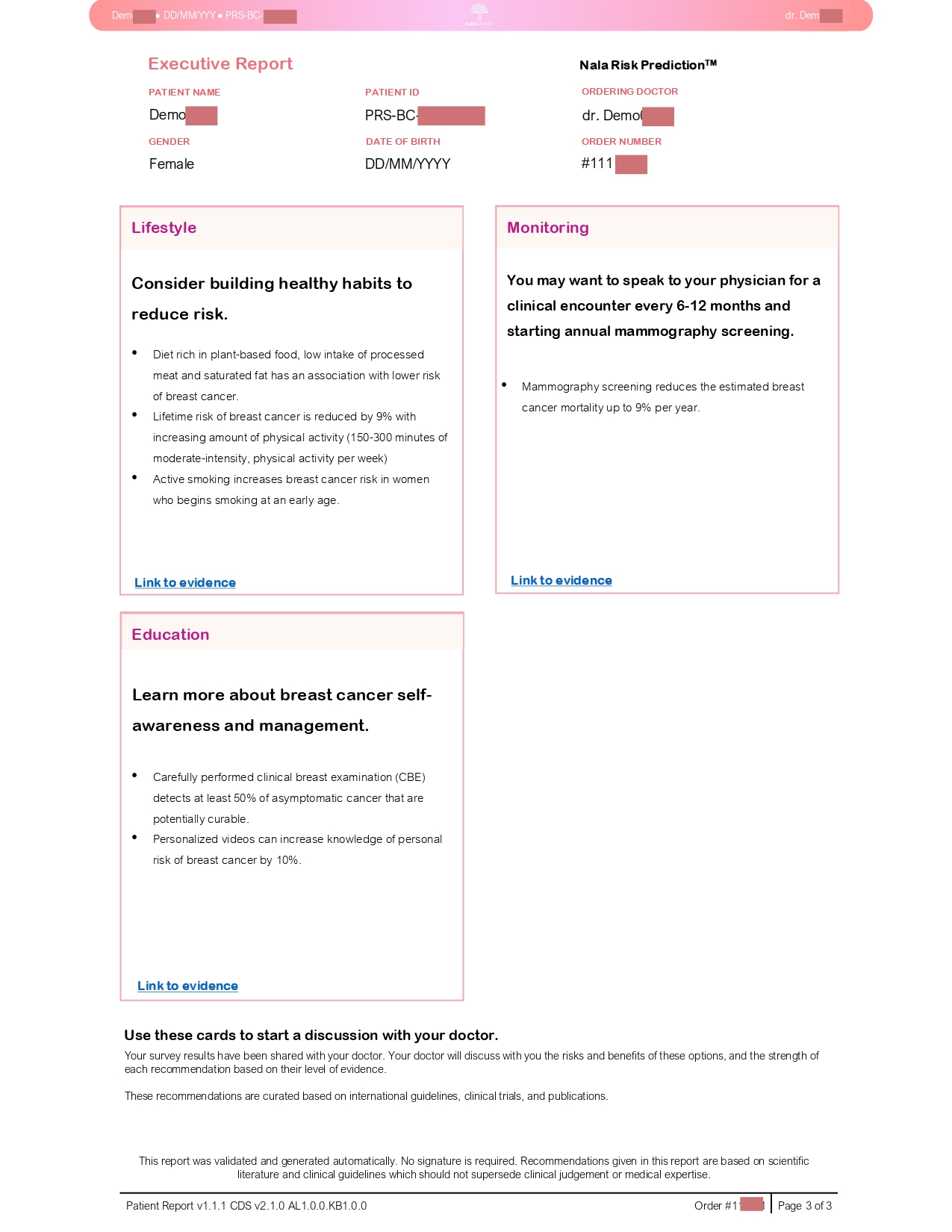

High Risk Report ([Link](https://nalagenetics.sharepoint.com/:b:/r/sites/clinical-studies/Shared%20Documents/General/%5BID%5D%20Clinical%20Studies%20Documents/PERCEPTION%20-%20Nalagenetics%20x%20MRCCC%20x%20SJH%20Initiatives%20-%20Breast%20Cancer%20RPS%20FGD/Dummy%20Reports/SG%20Dummy%20Report%20High%20Edited.pdf?csf=1&web=1&e=DOOQvE))
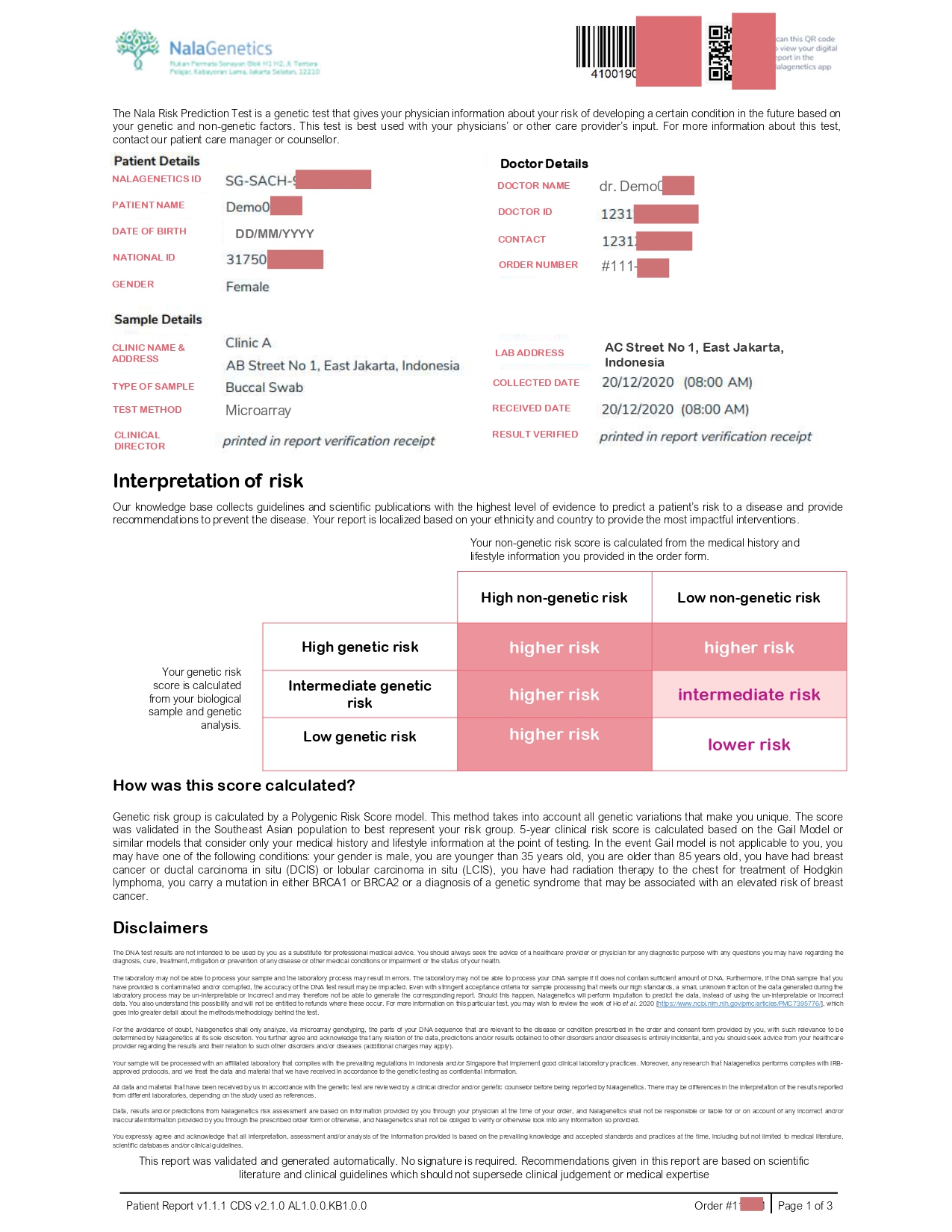

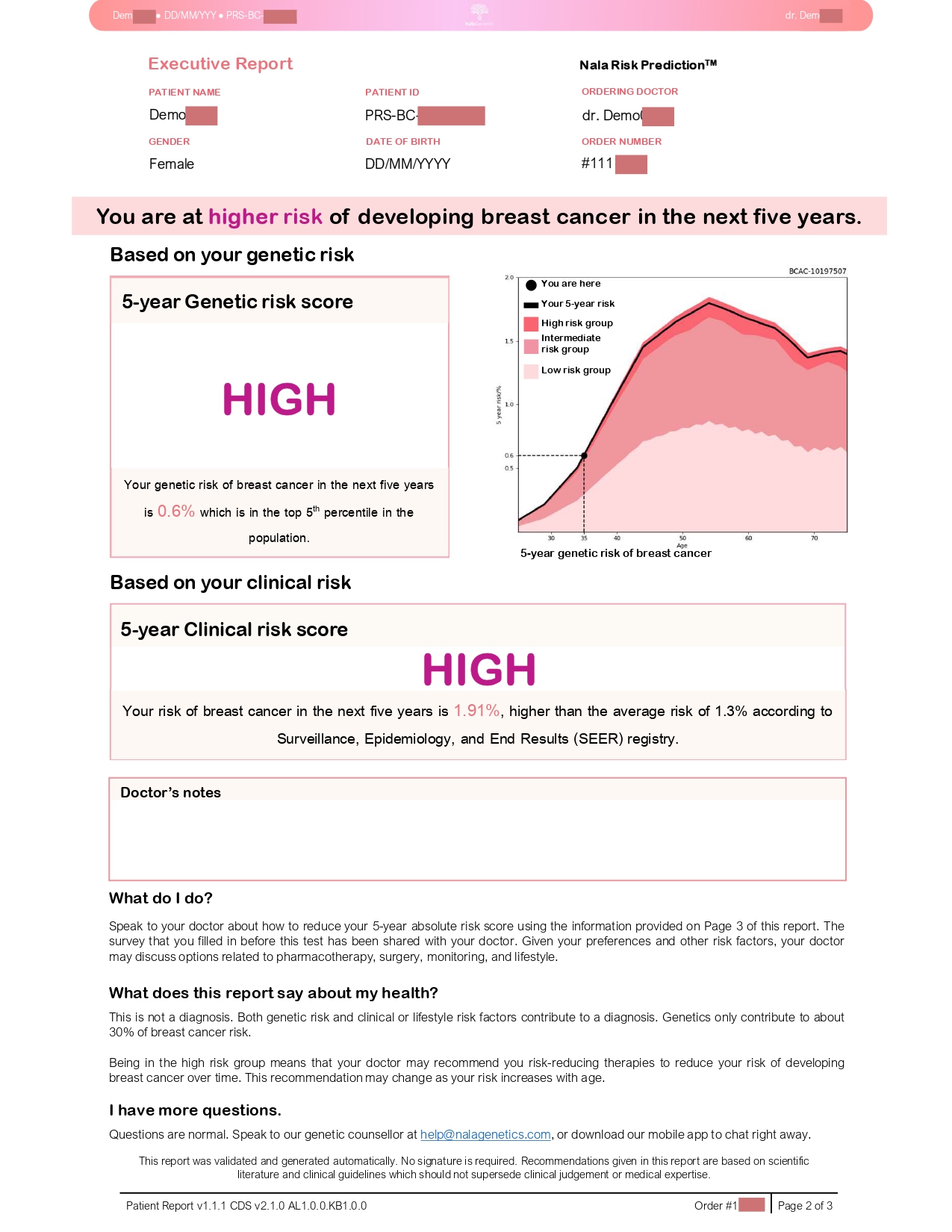

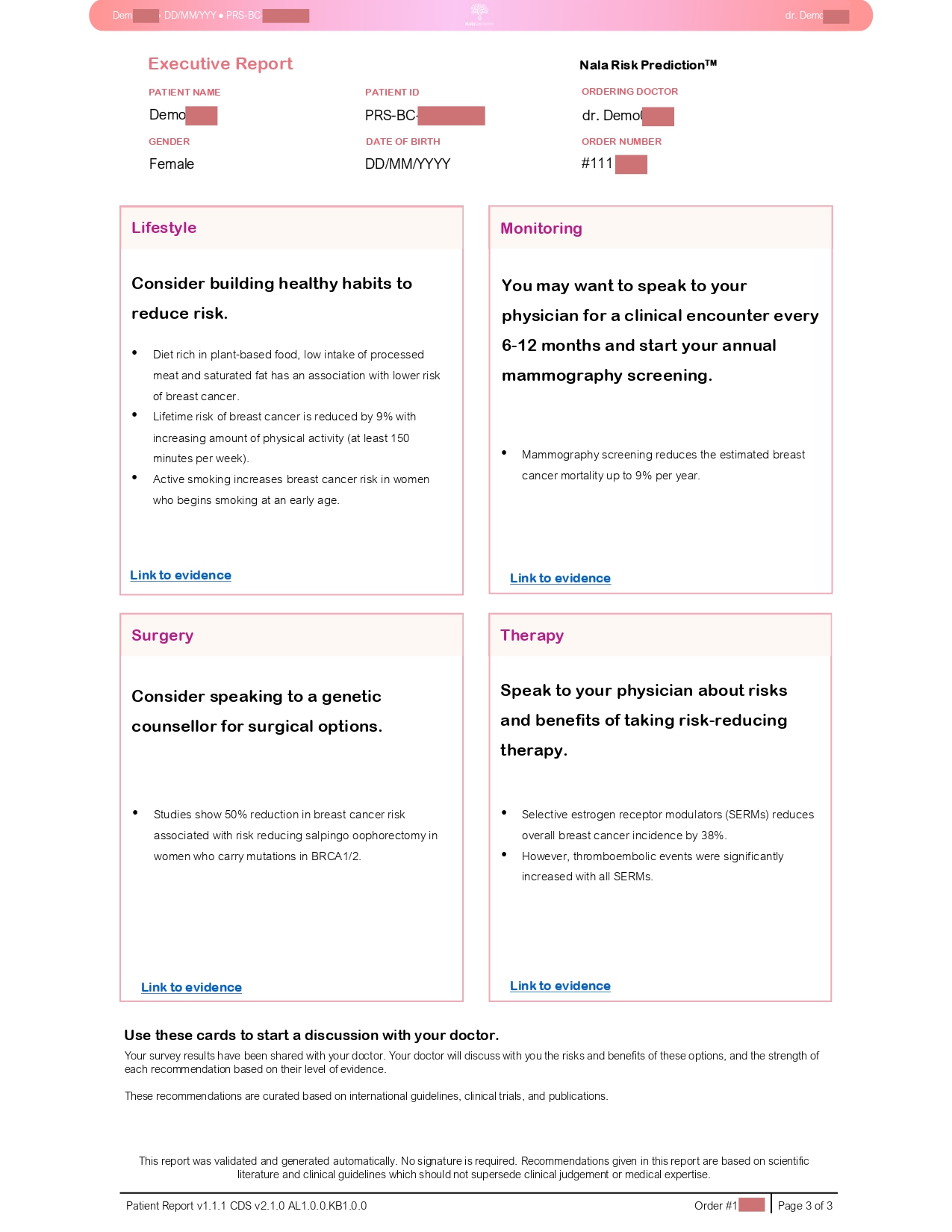

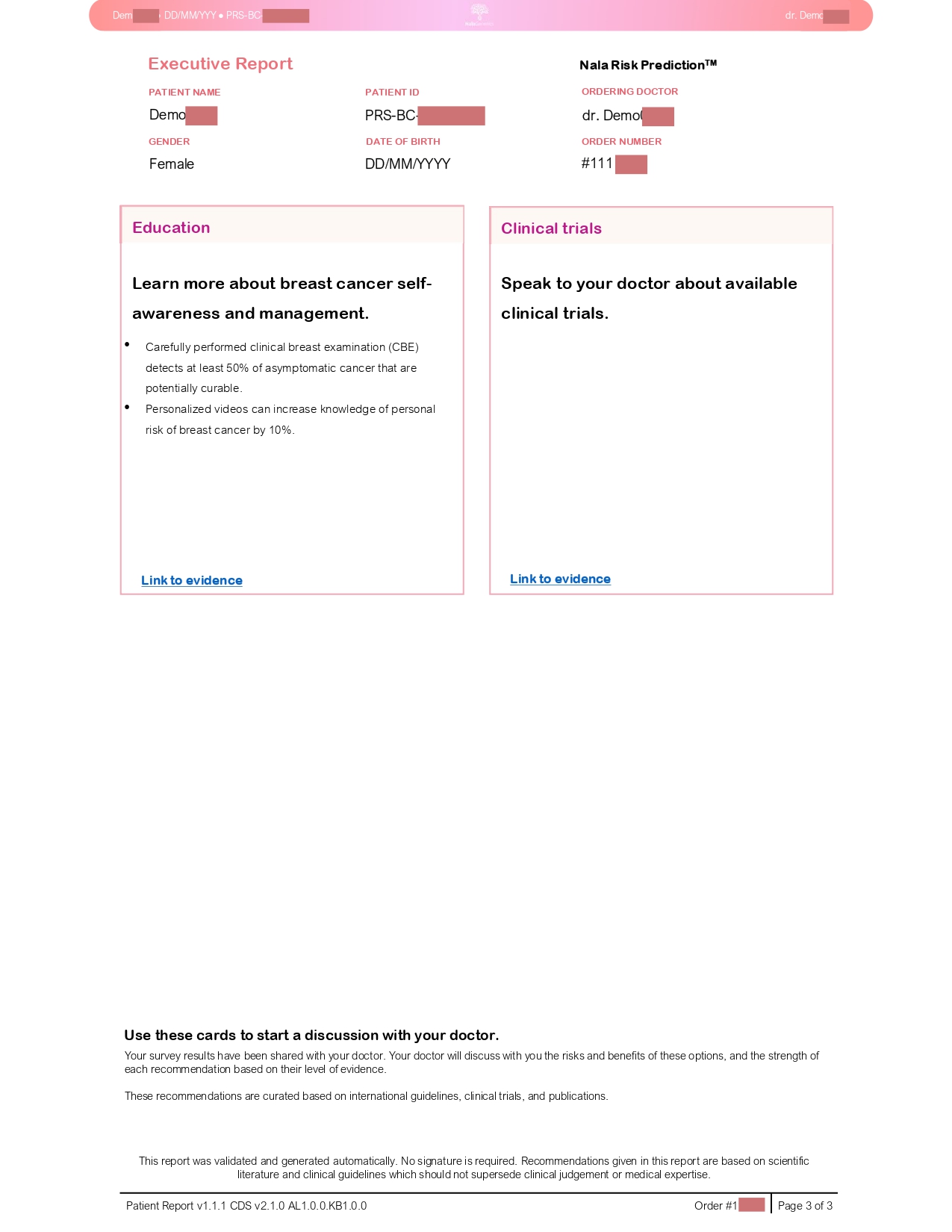
