## Supplementary material for "What do women want to see in a personalized breast cancer risk report? A qualitative study of Asian women of two countries": S3 File

**S3 File. FGD Question List**

1. This report expands on the previous findings but is calculated using the exact same methods. Which report would you prefer? (Applicable to Singapore FGD only)
2. Is the summary risk group clear to you?
3. Is the clinical risk calculation clear to you? Does the number help you understand your risk of breast cancer?
4. Is the genetic risk calculation clear to you? Does the number help you understand your risk of breast cancer?
5. What do you think about the recommendations listed? Do you think they are sufficient?
6. What questions would you ask your physician about each of the recommendations?
7. Imagine this report belongs to you. Upon receiving a "low/high risk" result, how would you feel?
8. Are there any other parties you would like to discuss these questions with?
9. How will this report change your relationship with your physician?
10. How will this report change your outlook about your health?
11. Would you want to see similar reports for other types of disease/ health screening?
12. If you want to similar reports for other types of disease/ health screening, do you have any specific diseases you are interested in?
13. What other changes can be made to improve your understanding of this report?
