## Supplementary figures and images for "What do women want to see in a personalized breast cancer risk report? A qualitative study of Asian women of two countries"

### S4 File

**S4 File. Simplified Report Prototype**

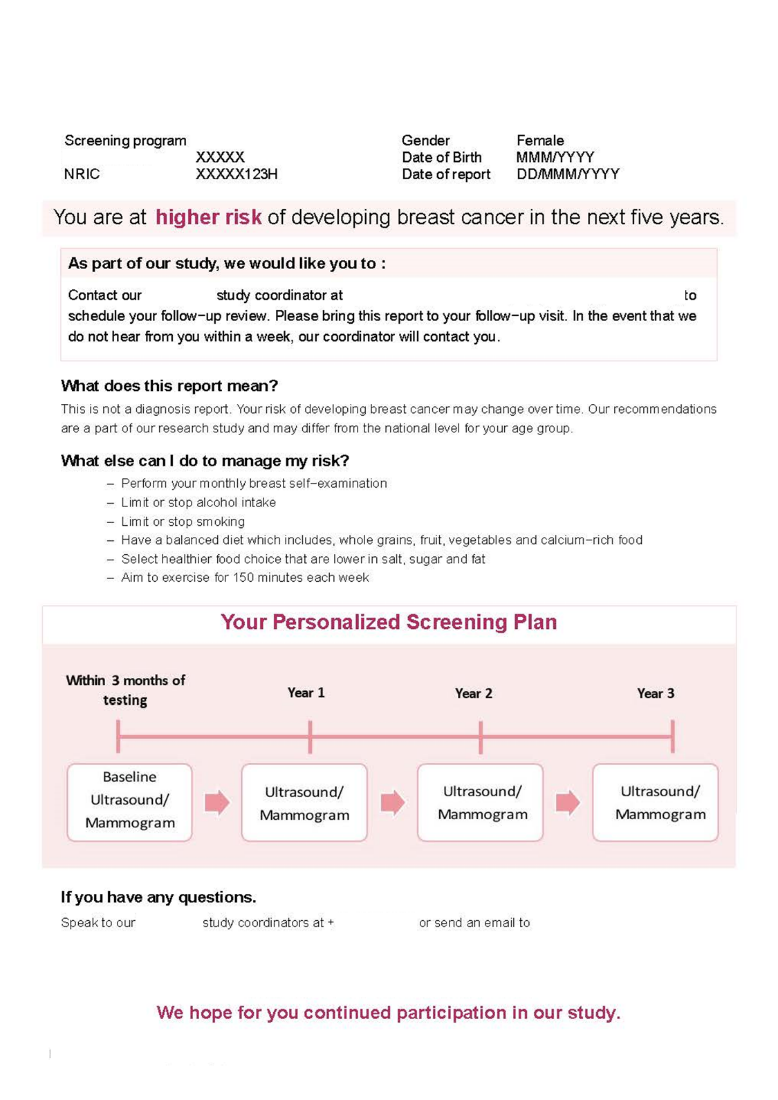
